# Hearing aids reduce daily-life fatigue and increase social activity: a longitudinal study

**DOI:** 10.1101/2021.05.05.21255749

**Authors:** Jack A. Holman, Avril Drummond, Graham Naylor

**Affiliations:** Hearing Sciences (Scottish Section), Mental Health and Clinical Neurosciences, School of Medicine, University of Nottingham, Glasgow, United Kingdom; School of Health Sciences, University of Nottingham, Nottingham, United Kingdom

**Author notes:** Correspondence: Jack A. Holman, Level 3 New Lister Building, Glasgow Royal Infirmary, 16 Alexandra Parade, Glasgow, G31 2ER.

**Keywords:** hearing loss, hearing handicap, listening effort, listening fatigue

## Abstract

People with hearing loss experience fatigue, and it is unknown whether this is alleviated by treatment with hearing aids. The objective of this study was to address this issue, and to investigate the possible concomitant effect of hearing-aid fitting on activity levels. An intervention group (n=53) who were due to be fitted with their first ever hearing aid(s) and a control group (n=53) who had hearing loss but no change in hearing aid status completed a battery of self-report outcome measures four times: once before fitting, and at two weeks, three months and six months post fitting. Self-report outcome measures at each assessment captured fatigue, listening effort, hearing handicap, auditory lifestyle, social participation restrictions and work, social and physical activity levels. Hearing-aid fitting led to a significant reduction in listening-related fatigue, but not general fatigue, in the intervention group compared to the control group. Additionally, social activity level increased and social participation restriction decreased significantly after hearing aid fitting in the intervention group compared to the control group. No significant interaction was found between working status and change in listening-related fatigue score. This study is the first to make longitudinal measurement of fatigue before and after first-ever hearing aid fitting and to identify an increase in social activity level after hearing aid fitting. These findings have important implications for future research and the clinical practice of hearing aid fitting.

## Introduction

Hearing loss can affect people’s lives beyond their difficulty hearing (Heffernan et al., 2016). One effect which has attracted increased interest in recent years is the potential additional fatigue experienced in everyday life by people with a hearing loss. While the experience of having fatigue may in itself be negative, fatigue can also negatively impact other health related outcomes such as activity and quality of life (Flensner et al., 2013). Thus, it is important to investigate ways of limiting the development and impact of fatigue in this population.

The basic and most intuitive theory of listening-related fatigue states that the cognitive effort required in challenging listening situations constitutes a drain on finite cognitive resources, resulting in fatigue (Hornsby et al., 2016). While this does seem to be a main cause of fatigue for people with a hearing loss (who experience challenging listening relatively frequently), the development of fatigue in some has also been linked to the negative emotions associated with having a hearing loss (Holman et al., 2019). A systematic review found some evidence that people with a hearing loss experience greater fatigue than those without (Holman, Drummond, et al., 2021). However, not all research has identified increased levels of fatigue in those with a hearing loss. Using Ecological Momentary Assessment, Burke and Naylor (2020) found no difference in general daily-life fatigue between people with hearing losses, as determined by audiometry, and those without. However, the control group in that study did have hearing difficulties such as tinnitus.

Logically, an increase in audibility through the fitting of a hearing device should reduce the listening effort required in any given situation, and in turn reduce fatigue. Evidence regarding the impact of hearing aid fitting (as opposed to cochlear implant fitting) on fatigue has focused to date mainly on the cross-sectional study of people wearing and not wearing hearing devices (Alhanbali et al., 2017; Bisgaard & Ruf, 2017). Such research has provided mixed evidence and, by virtue of using self-report questionnaires, invariably measures long-term fatigue. However, Hornsby (2013) used a crossover study design to measure transient fatigue and listening effort, and identified an objective benefit of hearing aid fitting. The lack of subjective benefit found by that study suggests that subjective and objective measures of transient fatigue may not tap into the same phenomenon. Regarding long-term fatigue, it is possible that general fatigue questionnaires may not consistently detect any beneficial impact of hearing device fitting on fatigue, due to a lack of sensitivity.

Listening-related fatigue affects people in different ways, and may also be managed differently from person to person (Holman et al., 2019; Davis et al., 2020). As listening-related fatigue is generally contingent on engagement in conversational activity, it has been postulated that activity levels and listening-related fatigue are related and may affect well-being (Holman, Hornsby, et al., 2021). It is thus possible that more conversationally active lifestyles would lead to greater instances of listening-related fatigue. Additionally, there is cross-sectional evidence that hearing aid use is linked to increased social activity levels (Fisher et al., 2015; Lee & Noh, 2015; Sawyer et al., 2019). Therefore, the impact of hearing aid fitting on listening-related fatigue may be connected to daily-life activity, and the impact of hearing aid fitting on well-being would be dependent on what, if any, changes there were in both fatigue and activity. Examining interactions between fatigue and activity has not been a feature of quantitative research in the field, which may account for discrepancies between studies.

Longitudinal assessment of fatigue before and after hearing aid fitting has not previously been reported. Therefore, we conducted a longitudinal study to address the following research questions:

(Q1) Does first-ever hearing aid fitting have an impact on fatigue?

We hypothesised that fatigue would reduce post hearing aid fitting.

(Q2) What variables may influence the impact of hearing aid fitting on fatigue?

Demographic, hearing and lifestyle factors could be associated with the impact of hearing aid fitting on fatigue. It was hypothesised that age, gender, hearing loss, hearing handicap, social activity level, work activity and need for cognition (inclination towards effortful cognitive activity) would be associated with fatigue scores. It was hypothesised that listening effort, being functionally linked to fatigue, would correlate temporally with fatigue.

(Q3) Do levels of social or physical activity change after hearing aid fitting, and is this related to fatigue?

We hypothesised that social and physical activity would rise after hearing aid fitting. Associated variables of interest for social activity were age, gender, hearing handicap, auditory lifestyle demand and need for cognition. Associated variables of interest for physical activity were age, gender and hearing handicap. We hypothesised that social activity would rise for participants who experienced a reduction in fatigue.

## Materials and Methods

This research received ethical approval from the West of Scotland Research Ethics Committee (18/WS/0030) and the NHS R&D (GN18EN073).

### Design

Fatigue was measured before, and at three timepoints after first-ever hearing aid fitting in an intervention group. A control group also completed the same measurement regime.

Additionally, several variables potentially related to fatigue were measured in order to assess potential mediating or associated factors, including accounting for individual differences at assessment 1 (baseline).

### Participants and Recruitment

Participants were between 18 and 75 years old. For inclusion in the intervention group participants had to have a self-reported hearing loss, be receiving their first ever hearing aid(s) and not be attending the audiology clinic with a primary complaint of tinnitus or vestibular issues. For inclusion in the control group participants had to have a self-reported hearing loss and have experienced no change in hearing aid status for at least one year. The control group was broadly matched with the intervention group based on age, gender and hearing loss (see Table 1).

**Table 1:** Group hearing and demographic characteristics

|  | **INTERVENTION** | **CONTROL** |
| --- | --- | --- |
| **N** | 53 | 53 |
| **FEMALE** | 20 | 29 |
| **AGE RANGE (MEAN, SD)** | 46-75 (61.4, 7.6) | 20-73 (66, 11.9) |
| **BE4FA RANGE (MEAN, SD)** | 10-88.75 (32.3, 13) | 7.5-107.5 (35.8, 19.9) |
| **WE4FA RANGE (MEAN, SD)** | 22.5-90 (40.1, 14.5) | 15-110 (44.4, 23.5) |
| **0HA** | 0 | 29 |
| **1HA** | 28 | 10 |
| **2HA** | 25 | 14 |
| **TINNITUS** | 18 | 24 |
| **EMPLOYED** | 14 | 12 |

BE4FA = Better ear four frequency average hearing loss (dBHL); WE4FA = Worse ear four frequency average hearing loss (dBHL); HA = Hearing aid; SD = Standard deviation

Participants for the intervention group were recruited from the waiting list for audiological evaluation at the Glasgow Royal Infirmary audiology department, except for two participants recruited through word of mouth. The first (baseline) assessment was conducted before each participant’s audiological screening, and for this reason not all potential intervention group participants ended up receiving hearing aids. Those who were not fitted with a hearing aid became part of the control group. The remaining participants in the control group were selected from the Hearing Sciences – Scottish Section participant pool. The stability of hearing aid status was monitored across the study to ensure participants remained eligible for their designated group.

A minimum sample size of 90 participants (45 per group) was estimated based on between-group analysis of covariance (ANCOVA), with a power of 80% (β=0.2), capable of detecting an effect size of 0.3 and a type one error rate alpha of 0.05. The power calculation was conducted using G* power calculator version 3.1. The sample size calculation was based on the Fatigue Assessment Scale (FAS) as this was the questionnaire used in the most relevant previous research (Alhanbali et al., 2017).

In total 129 participants completed at least one assessment, and 106 (49 female) completed all four assessments. Altogether 89 participants were recruited for potential inclusion in the intervention group. Seventeen of those participants did not complete all four assessments, dropping out for reasons such as personal circumstances, failure to respond to communications, and failure to meet the inclusion criteria that was not initially detected. Seven participants were offered a hearing aid at their clinical appointment, but chose not to be fitted, and twelve were told that a hearing aid would not be appropriate. Table 1 shows the hearing and demographic characteristics of participants. Four participants from the control group and 10 from the intervention group had worse-ear four-frequency averages (WE4FA) of <25 dB HL (500 Hz, 1, 2, and 4 kHz), which in some circumstances is categorised as no hearing loss. However, all these participants had a self-reported hearing loss based on a 5-point scale from no hearing loss to profound hearing loss.

### Procedure

Data collection ran from March 2018 until December 2019. Each participant made four visits to the department, with assessments spaced three months apart for the control group. For the intervention group there was one assessment before hearing aid fitting, and then two weeks, three months and six months post fitting. These time points were chosen as the first few weeks after a hearing aid fitting involve potentially troublesome sensory and psychosocial adjustments (Dawes et al., 2014), whereas by three months post fitting substantial performance improvements are regularly shown (Munro & Lutman, 2003; Munro, 2008). The only time interval that varied depending on availability of audiological screening appointments was from assessment 1 (baseline) to assessment 2, with some being three months apart and others being longer. The timeline of sessions and participant numbers is shown in figure 1.

**Figure 1:**
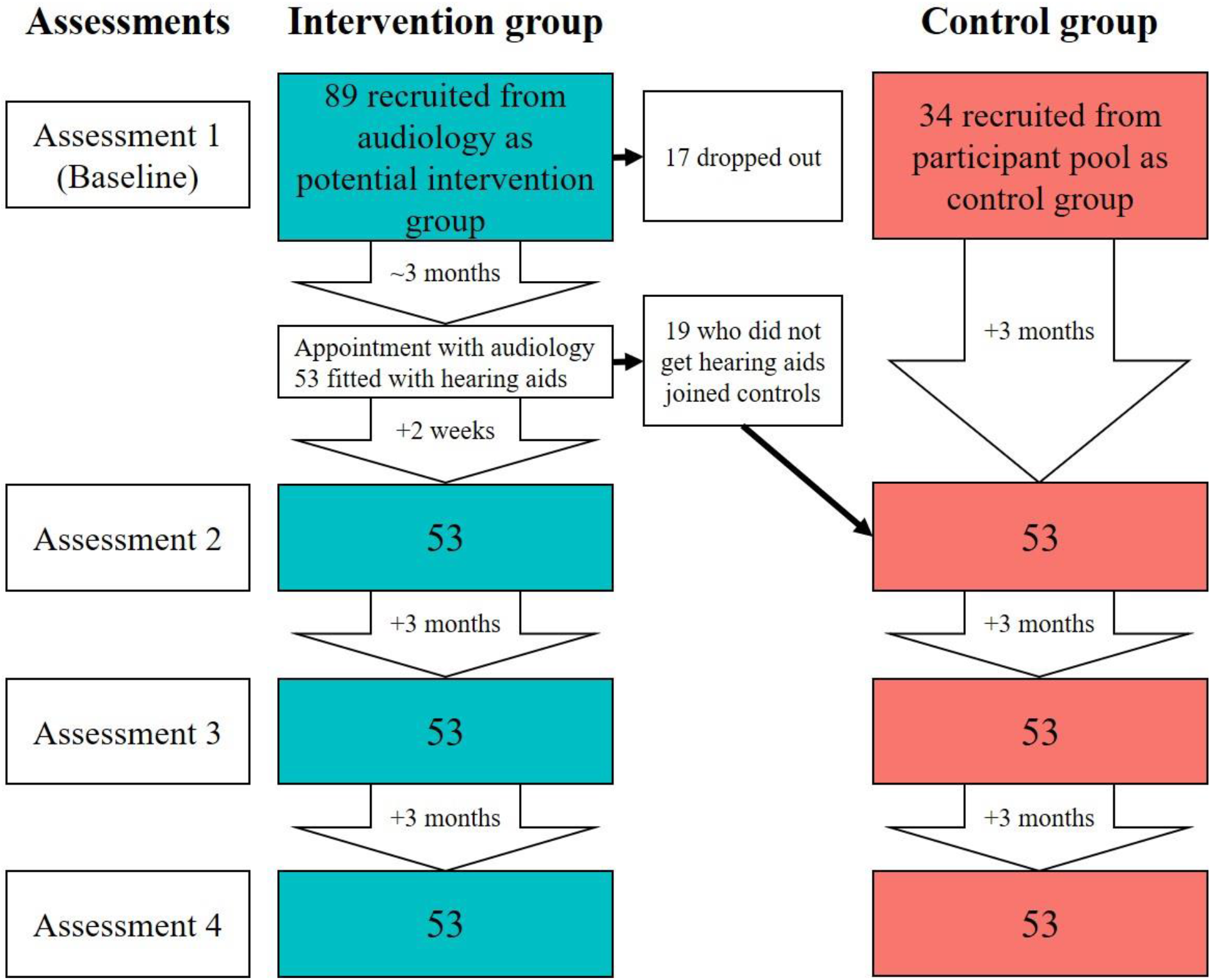
Timeline of assessments and participant numbers for control and intervention groups. The assessments are displayed on the left of the figure, with the timing of assessments displayed in downward arrows for each group. Dropouts occurred throughout the course of the study but are listed at baseline.

During assessment 1, which took approximately one hour, participants provided informed consent and were then asked about their hearing history (including hearing aid use and tinnitus), underwent ear examination and audiometric assessment, and completed research questionnaires. At subsequent assessments, participants completed the same questionnaires apart from the ‘Need for Cognition Scale’, and there was no repeat of the pure tone audiometry or history taking. Continued hearing aid use was ascertained at each assessment. While some participants rarely used their new hearing aids, no participants gave up on their hearing aids entirely. At the final assessment participants were debriefed and offered the opportunity to ask additional questions about the study. Appointments were arranged by either phone or email, depending on the participant’s preference, and appointment reminders were also provided in the same way. Participants received £50 compensation in total; £10 at each of the first three visits, and £20 at the final visit.

### Outcome measures

The outcome measures used in the study were thirteen self-report questionnaires which addressed fatigue, listening effort, hearing handicap, activity (social, work and physical), social participation restrictions, auditory lifestyle, and need for cognition. For the purposes of this study, participants were asked to respond with respect to their current level of hearing aid usage (i.e. in situations where they would be wearing their hearing aids, answering the corresponding question as though wearing hearing aids). Full questionnaire details are provided in supplementary digital content 1.

Three self-report questionnaires in total were utilised to measure fatigue. Two of the questionnaires assessed long-term fatigue. The Fatigue Assessment Scale (FAS) (Michielsen et al., 2003) is a widely used unidimensional general fatigue scale. The Multidimensional Fatigue Symptom Inventory – Short Form (MFSI) (Stein et al., 2004) measures the domains of general fatigue, physical fatigue, emotional fatigue, mental fatigue and vigour (Donovan et al., 2015). One self-report questionnaire assessed listening-related fatigue specifically. The Vanderbilt Fatigue Scale – Adult Hearing Loss (VFS-AHL) (Hornsby et al., 2021) comprises of both unidimensional and multidimensional scales. The multidimensional scales assessed in the current study were cognitive fatigue, emotional fatigue and social fatigue.

Listening effort was assessed using the Listening Effort Assessment Questionnaire (EAS) (Alhanbali et al., 2017) which measures the amount of effort participants use “listening in everyday life”. Hearing handicap was assessed using the Hearing Handicap Inventory for the Elderly/Adults (HHIE/A) (Ventry & Weinstein, 1982; Newman et al., 1990), the HHIE for people aged 65 and over; otherwise HHIA.

Social activity level was assessed using two self-report questionnaires. The Social Activity Log (SAL) (Syrjala et al., 2010) measures the quantity of social activity in the “past week” and “past month”. The Social Participation Questionnaire (SPQ) (Densley et al., 2013) measures the quantity of social activity undertaken in the “last twelve months”. Work activity was assessed using the “how do you spend your time?” section of the Craig Handicap Assessment and Reporting Technique (CHART) (Whiteneck et al., 1992). Physical activity in the “last seven days” was assessed using the International Physical Activity Questionnaire - Short (IPAQ) (Craig et al., 2003).

The experiential component of social activity was assessed using the Social Participation Restrictions Questionnaire (SPaRQ) (Heffernan et al., 2018; Heffernan et al., 2019) which consists of two scales assessing ‘social behaviours’ and ‘social perceptions’. Auditory lifestyle was assessed using the Auditory Lifestyle and Demand Questionnaire (ALDQ) (Gatehouse et al., 1999). The tendency for an individual to engage in and enjoy thinking was assessed using the Need for Cognition Scale (NFC) (Cacioppo & Petty, 1982).

### Statistical Analysis

Statistical analyses were carried out using R version 3.6.1 (R Core Team, 2019). For analyses of group means, analyses of covariance (ANCOVA) were conducted where appropriate. As the data were longitudinal and the individual trajectories were important, multilevel modelling (growth curve modelling specifically) was used as the primary approach to analysis. This was analysed using the nlme package (Pinheiro et al., 2019). The hierarchy in the dataset was repeated measures (level 1) nested within individual participants (level 2).

Associations between group (Intervention vs. Control) and outcome measure scores at baseline was assessed using independent-sample t-tests, or Mann-Whitney U tests. Group mean differences at each subsequent time point, controlling for baseline score, were analysed using ANCOVA or non-parametric equivalent. The sm package was used to analyse non parametric ANCOVA (Bowman & Azzalini, 2018). Separate multilevel growth models were built to assess the relationships between group and outcome measures across time for individuals, followed by assessment of the intervention group alone where relevant. Kendall rank correlation coefficients (rτ) were used to assess the relationships between magnitude of change in relevant variables.

## Results

In all following text, the pre-fitting assessment is termed ‘baseline’ with the post fitting assessments denoted as 2, 3 and 4. A Spearman rank correlation matrix of all baseline questionnaires is provided in supplementary digital content 2.

### Research Question 1: Does first-ever hearing aid fitting have an impact on fatigue?

Following ANCOVA analysis and after controlling for baseline scores, FAS scores were significantly different between groups (higher in controls) at assessment 2 (H= 2.82, p=.012), but not at assessments 3 or 4 (figure 2). There was no significant difference between groups at any of the three post fitting assessments for total MFSI score after controlling for baseline scores (figure 2). Unidimensional VFS-AHL scores, after controlling for baseline scores, were significantly lower (better) in the intervention group at all subsequent assessments (2: F(1, 103) = 16.33, p=<0.001; 3: F(1, 103) = 40, p=<0.001; 4: F(1, 103) = 39.8, p=<0.001) (figure 2).

**Figure 2:**
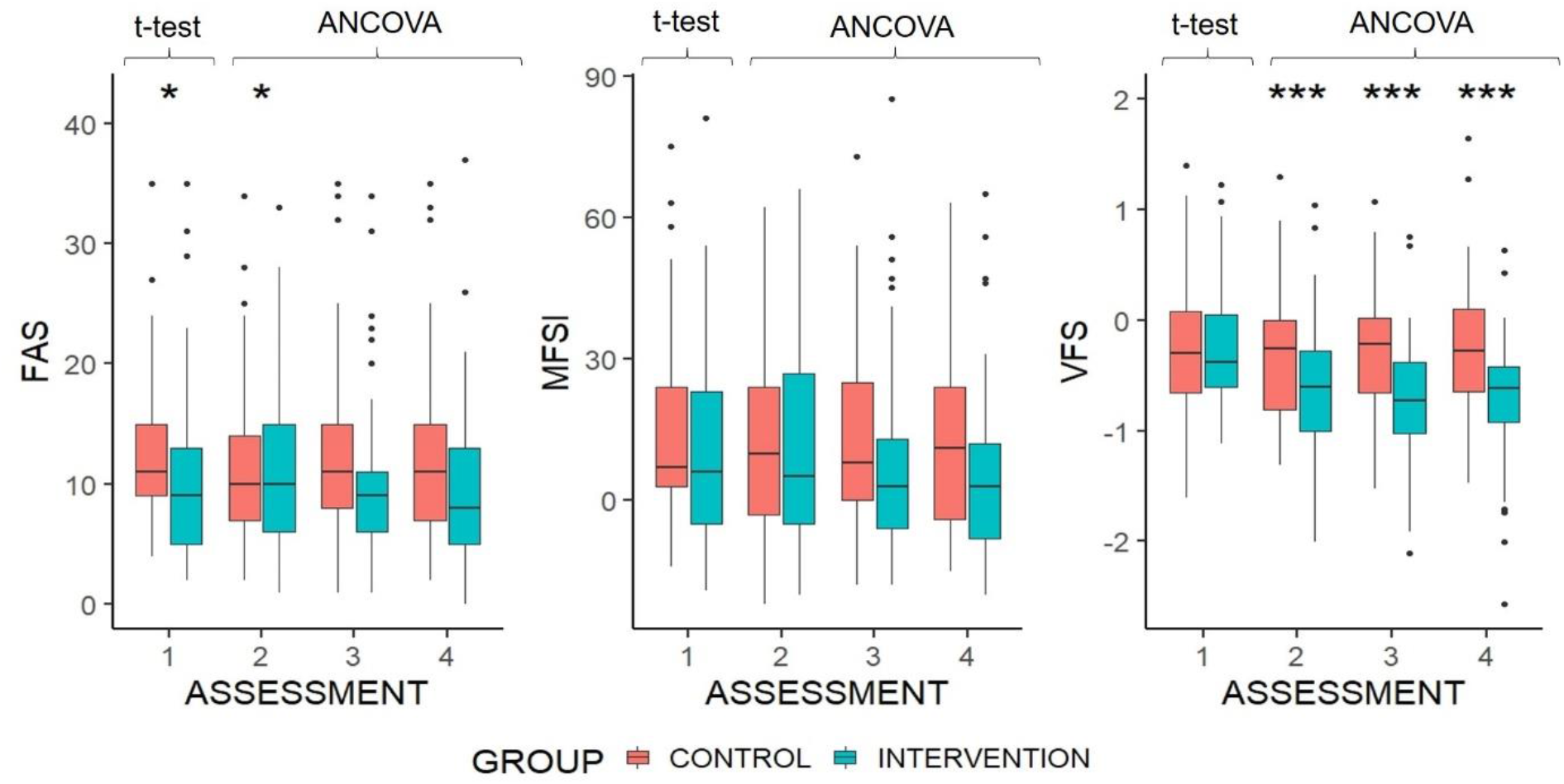
Boxplots of FAS, MFSI and VFS-AHL score by group and assessment. Group difference at baseline is based on independent samples t-test. Differences at each subsequent time point are based on ANCOVA, controlled for baseline scores. Higher scores indicate greater fatigue. *p < .05 **p < .01 ***p < .001 Upper and lower hinges represent 25th and 75th percentiles. Upper whisker is highest value within 1.5 x inter-quartile range, measured from the upper quartile. Lower whisker is the lowest value within 1.5 x inter-quartile range, measured from the lower quartile. Values beyond this are represented as outlier dots.

Analysis of the multidimensional scales of the MFSI, after controlling for baseline scores, found no significant differences between groups in any assessment for physical fatigue, mental fatigue or emotional fatigue. The vigour scale of the MFSI showed that after controlling for baseline scores, intervention group scores were significantly higher (better) than the control group at the final two assessments, (F(1, 103) = 4.56, p= .039; F(1, 103) = 6.19, p= .015). VFS-AHL scores (baseline corrected) for cognitive fatigue, emotional fatigue and social fatigue were lower in the intervention group than the control group at assessment 2, (F(1, 103) = 19.01, p= < .001; F(1, 103) = 11.25, p= .001; F(1, 103) = 13.74, p= < .001), assessment 3, (F(1, 103) = 41.12, p= < .001; F(1, 103) = 22.25, p= < .001; F(1, 103) = 36.69, p= < .001), and assessment 4, (F(1, 103) = 32.86, p= <.001; F(1, 103) = 35.67, p= < .001; F(1, 103) = 49.91, p= < .001). Based on these results, there does appear to be a beneficial impact of first-ever hearing aid fitting on fatigue (Q1).

Following on from the group level analysis, multilevel growth model analysis was conducted to assess fatigue at the participant level across both groups. Group and baseline HHIE/A score were significantly associated with VFS-AHL score, with fatigue increasing with HHIE/A score (table 2). Assessment number was not significantly associated with VFS-AHL score, but the interaction between group and assessment number was, suggesting that the difference between groups varied across assessments. There was no significant association between VFS-AHL score and age, sex, BE4FA, WE4FA, NFC score, or baseline SAL score.

**Table 2:** Effects of predictors on VFS-AHL score: all participants

| CONSTRUCT | B | SE | T |
| --- | --- | --- | --- |
| **GROUP** | -0.25 | 0.09 | -2.88** |
| **ASSESSMENT NUMBER** | -0.46 | 0.36 | 1.28 |
| **AGE (GMC)** | -0.004 | 0.004 | -0.9 |
| **SEX** | 0.13 | 0.082 | 1.59 |
| **BE4FA** | -0.001 | 0.004 | -0.29 |
| **WE4FA** | 0.002 | 0.004 | 0.53 |
| **BASELINE SAL** | 0.008 | 0.039 | 0.21 |
| **BASELINE HHIE/A** | 0.016 | 0.002 | 7.28*** |
| **NFC** | 0.001 | 0.003 | 7.28 |
| **GROUP X ASSESSMENT** | 1.2 | 0.51 | 2.37* |

b = unstandardized beta; SE = standard error; t test statistic; GMC = Grand mean centred; SAL = Social Activity Log; HHIE/A = Hearing Handicap Inventory for the Elderly/Adults; NFC = Need for Cognition Scale; VFS-AHL = Vanderbilt Fatigue Scale – Adult Hearing Loss
\*p < .05 \*\*p < .01 \*\*\*p < .001

### Research Question 2: What variables may influence the impact of hearing aid fitting on fatigue?

To further investigate the impact of hearing aid fitting on fatigue by breaking down the interaction term (group x assessment) and re-assessing the relationship between fatigue and associated variables, a new multilevel growth model was created for the intervention group only (table 3). The slopes did not vary significantly across participants, SD = 0.46 (95% CI: 0.36, 0.6), x^2^(2) = 3.92, p= .14, suggesting that participants in the intervention group tended to follow the same trend across time. A logarithmic function of time was found to be a better fit than linear, quadratic and cubic, SD = 0.4 (95% CI: 0.31, 0.5), x^2^(2) = 4.66, p= .03.

**Table 3:**
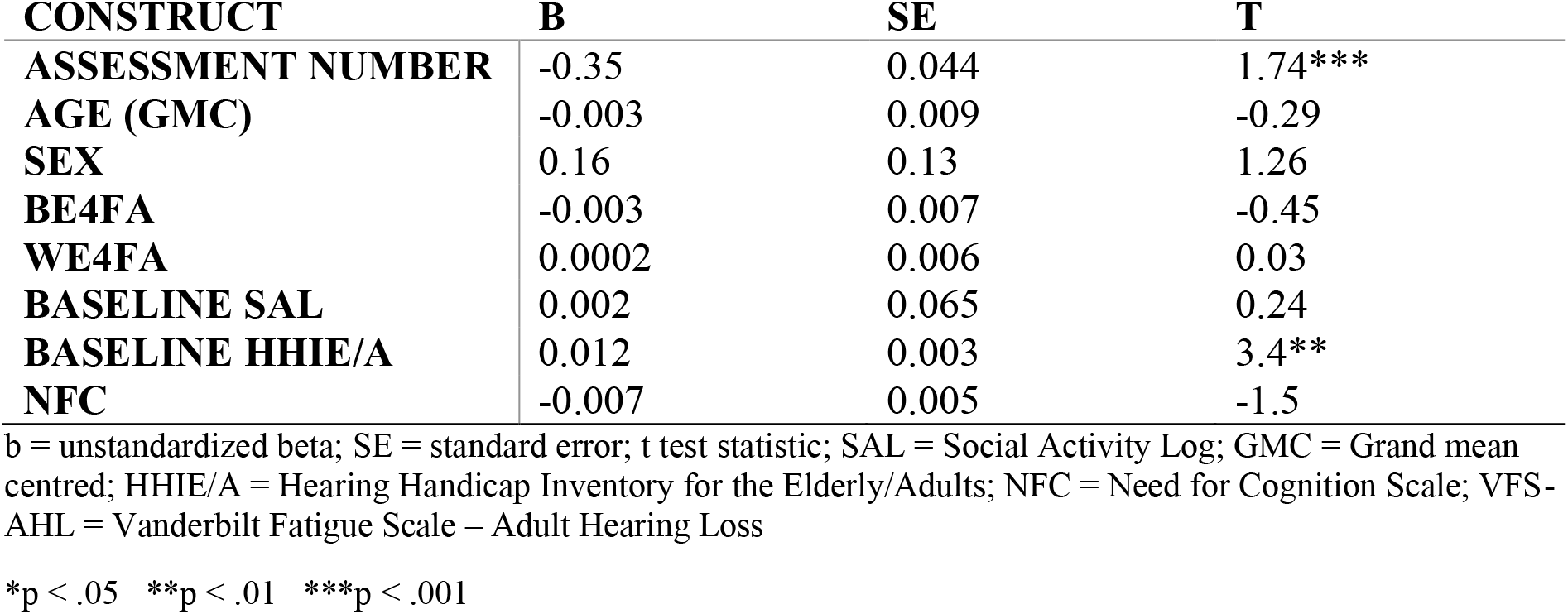
Effects of predictors on VFS-AHL score: intervention group

While the growth model analysis suggested a logarithmic trend of VFS-AHL over time, visualisation of the individual VFS-AHL trajectories highlights that individuals had a wide variety of score trajectories from baseline to assessment 4 (supplementary digital content 2).

Baseline HHIE/A was significantly and positively associated with VFS-AHL score in the intervention group, and assessment number was significantly and negatively associated with VFS-AHL score (table 3). There was no significant association between VFS-AHL score and age, sex, BE4FA, WE4FA, NFC score or baseline SAL score.

After controlling for baseline scores, listening effort significantly decreased in the intervention group post fitting compared to the control group, (EAS: F(1, 103) = 11.04, p= .0012; F(1, 103) = 32.92, p= < .0001; F(1, 103) = 22.46, p=<. 0001). In the intervention group, the change in VFS-AHL score from baseline was significantly correlated with the change in EAS score at all follow up assessments (2: rτ = 0.32, p = .0007; 3: rτ = 0.22, p = .02; 4: rτ = 0.42, p = < .0001).

#### Work Activity

Mean VFS-AHL scores for intervention group participants who work were higher at each assessment than for those who were not in work. However, this difference was not statistically significant at baseline, t=-0.57, p=.57. There was a significant difference when controlling for baseline scores at the second assessment, (F (1, 50) = 6.52, p = 0.014), but not at the final two post intervention assessments, (F (1, 50) = 0.62, p = 0.44; F (1, 50) = 3.9, p = 0.054). The lack of a consistent pattern suggests that post intervention VFS-AHL scores changed at a similar rate in both working and non-working participants.

In summary, hearing handicap at baseline is the only pre-fitting variable which was significantly associated with change in fatigue, and which therefore may influence the impact of hearing aid fitting on fatigue (Q2).

### Research Question 3: Do levels of social or physical activity change after hearing aid fitting, and is this related to fatigue?

To answer this compound question, we analysed the activity levels across time between the intervention and control groups, followed by correlation analysis of the temporal changes in activity and fatigue.

#### Social activity

ANCOVA analysis found that SAL scores, controlling for baseline score, were significantly higher in the intervention group than the control group at assessments 2, 3 and 4 (figure 3). However, the mean SAL score for the control group dropped from baseline to assessment 2, which may have exaggerated the effect of hearing aid fitting. SPQ scores were significantly higher in the intervention group at the final assessment, with control group scores steady across assessments (figure 3).

**Figure 3:**
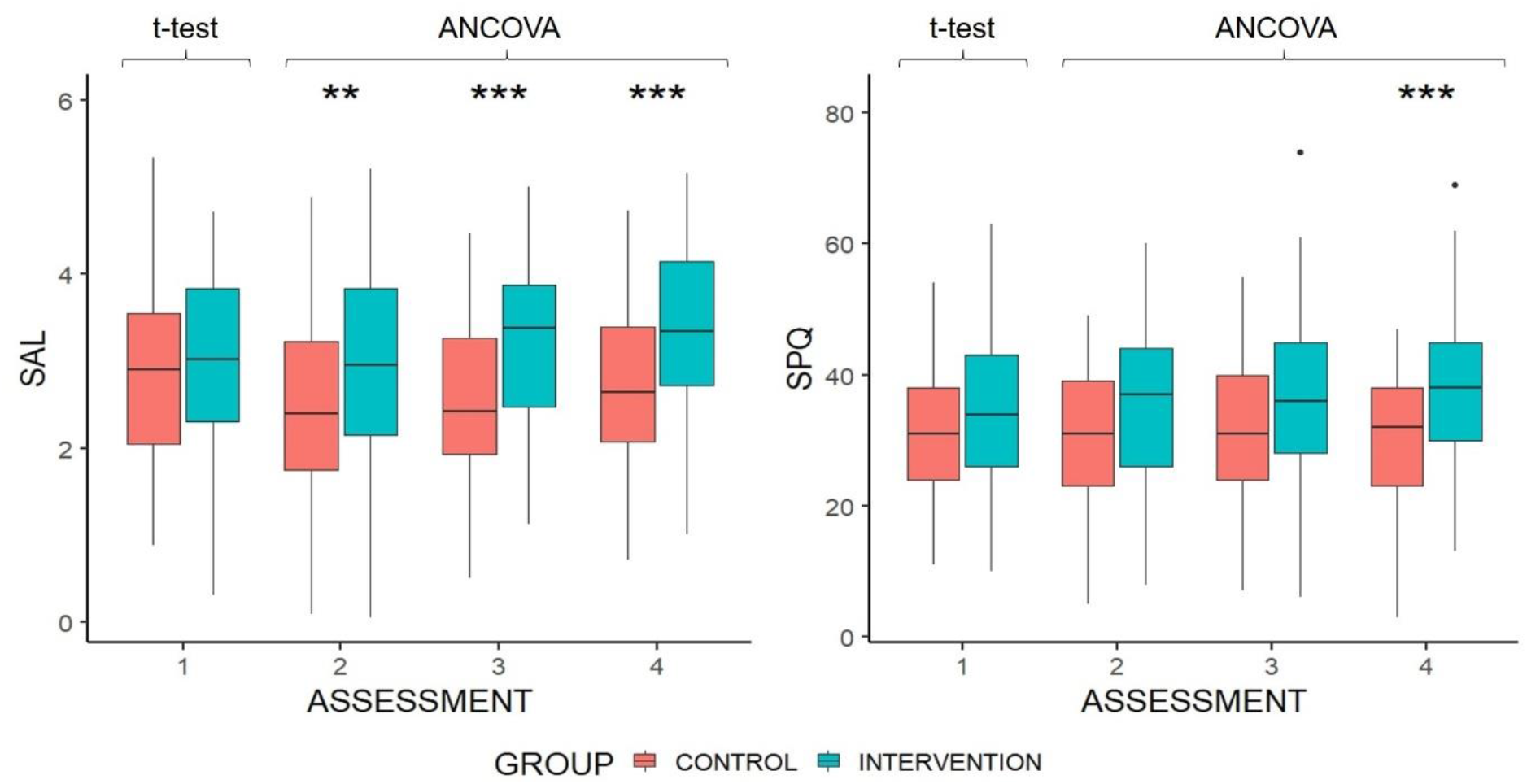
Boxplots of SAL & SPQ score by group and assessment. Group difference at baseline is based on independent samples t-test. Differences at each subsequent time point are controlled for baseline scores. Higher scores indicate greater social activity. *p < .05 **p < .01 ***p < .001 Upper and lower hinges represent 25th and 75th percentiles. Upper whisker is highest value within 1.5 x inter-quartile range, measured from the upper quartile. Lower whisker is the lowest value within 1.5 x inter-quartile range, measured from the lower quartile. Values beyond this are represented as outlier dots.

Group, assessment number, baseline ALDQ and NFC were significantly and positively associated with SAL score (table 4). Baseline HHIE/A was significantly and negatively associated with SAL score. There was no significant association between age, sex or the interaction of group and assessment and SAL score.

**Table 4:** Effects of predictors on SAL score: all participants

| CONSTRUCT | B | SE | T |
| --- | --- | --- | --- |
| ASSESSMENT NUMBER | 2.4 | 0.79 | 3.04** |
| GROUP | 0.41 | 0.15 | 2.67** |
| AGE (GMC) | 0.013 | 0.008 | 1.64 |
| SEX | -0.14 | 0.16 | -0.87 |
| BASELINE ALDQ | 0.027 | 0.006 | 4.7*** |
| BASELINE HHIE/A | -0.012 | 0.004 | -3.04** |
| NFC | 0.013 | 0.006 | 2.26* |
| GROUP X ASSESSMENT | -1.9 | 1.11 | -0.87 |

b = unstandardized beta; SE = standard error; t test statistic; GMC = Grand mean centred; ALDQ = Auditory Lifestyle Demand Questionnaire; HHIE/A = Hearing Handicap Inventory for the Elderly/Adults; NFC = Need for Cognition Scale
\*p < .05 \*\*p < .01 \*\*\*p < .001

As group was significantly associated with SAL score, further analysis was conducted on the intervention group alone. For this group, assessment and baseline ALDQ score were significantly and positively associated with SAL score (table 5). Baseline HHIE/A score was significantly and negatively associated with SAL score. There was no significant association between SAL score in the intervention group and age, sex, or NFC.

**Table 5:** Effects of predictors on SAL score: intervention group

| CONSTRUCT | B | SE | T |
| --- | --- | --- | --- |
| ASSESSMENT NUMBER | 0.15 | 0.033 | 4.69*** |
| AGE (GMC) | -0.008 | 0.014 | -0.53 |
| SEX | 0.015 | 0.22 | 0.07 |
| BASELINE ALDQ | 0.029 | 0.008 | 3.69*** |
| BASELINE HHIE/A | -0.02 | 0.005 | 3.85*** |
| NFC | 0.01 | 0.008 | 1.3 |

b = unstandardized beta; SE = standard error; t test statistic; GMC = Grand mean centred; ALDQ = Auditory Lifestyle Demand Questionnaire; HHIE/A = Hearing Handicap Inventory for the Elderly/Adults; NFC = Need for Cognition Scale
\*p < .05 \*\*p < .01 \*\*\*p < .001

Given that social activity increased significantly after hearing aid fitting, the change in social participation restriction was also investigated. It was found that the social benefit subscale of the SPARQ reduced (improved) significantly more in the intervention group than the control group when controlling for baseline scores at each assessment after hearing aid fitting, (F (1, 103) = 8.2, p = .005; F (1, 103) = 21.5, p = <.0001; F (1, 103) = 17.26, p = <.0001). The social perception subscale also reduced (improved) significantly in the intervention group at each post intervention assessment, (F (1, 103) = 4.16, p = .044; F (1, 103) =25.89, p = <.0001; F (1, 103) = 19.35, p = <.0001).

For participants in both groups, the changes in VFS-AHL and SAL scores from baseline to assessment 2 and 3 were not significantly correlated, but at assessment 4 the changes from baseline were significantly correlated, (rτ = -0.17, p = .009). However, when the intervention group alone was analysed, the correlation became marginal, (rτ = -0.18, p = .064). Therefore, social activity level increased with hearing aid fitting, but there is no evidence that decreasing fatigue is associated with increasing social activity (Q3).

#### Physical Activity

There was no mean difference of IPAQ scores between groups at baseline, U=1383, p = .89. There was also no significant difference when controlling for baseline scores at any of the post intervention assessments, F (1, 103) = 0.45, p = 0.5; F (1,103) = 0.58, p = .45; F (1, 103) = 1.05, p = .31. Therefore, there is no evidence of physical activity being affected by hearing aid fitting.

## Discussion

This study was designed to examine the impact of hearing aid fitting on fatigue, and variables related to changes in fatigue (research question Q1), whether activity changed after hearing aid fitting (Q2) and whether this had any link to fatigue (Q3). The two general fatigue questionnaires (FAS & MFSI) showed no difference between groups post fitting. This finding supports the results of Alhanbali et al. (2017), where FAS scores were not significantly different for hearing impaired groups with or without hearing aids. While cross-sectional research using another long-term general fatigue questionnaire has previously found a significant effect (Bisgaard & Ruf, 2017), the majority of support for a beneficial impact of hearing device fitting on long-term general fatigue has come from prospective non-randomised controlled trials of cochlear implants (Chung et al., 2012; Harkonen et al., 2015b, 2015a). This might suggest that the intervention in severe-profound hearing losses would result in a greater benefit for long-term general fatigue. It is also important to distinguish the difference between hearing aid fitting and hearing aid use. Discrepancies between the amount of time hearing devices were used may account for some differences in study results.

Due to the very recent development of the VFS-AHL scale, our finding of a significant impact of hearing aid fitting on VFS-AHL scores at both the group and individual levels stands alone, to date. The VFS-AHL measures listening-related fatigue, and as such seems more sensitive to the fatigue experienced by people with a hearing loss than general fatigue questionnaires, despite scores on all three fatigue questionnaires being significantly positively correlated at assessment 1. The VFS-AHL does not measure transient fatigue explicitly, but it does pose hypothetical situational questions. As a result, this study can offer no further insights into the distinct impacts of hearing aid fitting on transient and long-term fatigue. With the exception of the MFSI vigour subscale, the scores on all of the constituent subscales of the MFSI and the VFS-AHL followed the same pattern as the total scales. MFSI subscale scores were not significantly different between groups across time points, whereas VFS-AHL subscale score changes were significantly different between groups at each time point post fitting. These results could mean either that hearing aid fitting impacts all dimensions of listening-related fatigue similarly, or that people tend to report subjective fatigue as unidimensional, as has been previously suggested (Michielson et al., 2004). The lack of correlation between baseline hearing handicap and MFSI vigour does not support the previous finding of correlation between HHIE and the vigour subscale from the Profile of Mood States (Hornsby & Kipp, 2016). However, the significant improvement at the final two assessments in the intervention group suggests that vigour may play a role in listening-related fatigue.

Although a reduction in listening-related fatigue is clearly to be welcomed, it is unclear whether the change is of clinical significance. Median VFS-AHL scores were negative for both groups at baseline (on a scale of roughly 2 to -2), which suggests that listening-related fatigue was not high for most participants, and therefore may not have been problematic. It is currently not possible for us to report VFS-AHL data to establish whether the scores were normal for a population with audiometrically determined hearing loss, or how they compare to normal hearing norms. However, previous research has established that the threshold of minimally important difference for changes in health-related quality of life is approximately half a population standard deviation (Norman et al., 2003). At baseline, half a standard deviation in VFS-AHL score for the intervention group was 0.27. The change in mean VFS-AHL from baseline to the final assessment was -0.47, suggesting that the change in listening-related fatigue was indeed greater than the minimally important difference. For long-term general fatigue, FAS median scores calculated as a percentage were 22.5 % in the intervention group and 27.5% in the control group at assessment 1. This is similar to reports from previous research where the adjusted median FAS scores for three separate hearing loss groups were 22.5%, 22.5% and 22% (Alhanbali et al., 2017). This indicates that long-term general fatigue in the participant sample was at expected levels.

In line with Alhanbali and colleagues (2018), hearing handicap correlated with fatigue. The HHIE/A was the only questionnaire to correlate significantly with all three fatigue questionnaires at baseline, and significantly associated with VFS-AHL scores in the multilevel growth models. The absence of any relationship between fatigue and audiometric hearing loss reported by Alhanbali was also reflected in the current study. However, while Alhanbali et al. (2017) reported a significant relationship between FAS and EAS scores, here EAS was significantly correlated with VFS-AHL score. The finding that the change in VFS, EAS and HHIE/A scores correlated across time in the intervention group suggests that listening-related fatigue, hearing handicap and listening effort may be impacted in similar ways by hearing aid fitting. It is possible that the change in listening effort could have driven the change in fatigue. However, no causal relationship between the changes can be ascertained here.

Previous cross-sectional studies suggest that hearing device fitting has a positive effect on social activity (Fischer et al., 2015; Sawyer et al., 2019). This was supported by our longitudinal data, which is an important finding in its own right. The multilevel growth model analysis accounted for a decrease in SAL score in the control group, which, given that such a drop has no apparent plausible cause, may have skewed results at assessments 2 and 3. The differing time intervals for the measurement of social activity in the SAL (over the past month), and SPQ (over the past year), is reflected in the results, as significant improvements in the intervention group compared to the control group for the SAL were seen by assessment 2, whereas improvements in SPQ score were evident only at the final assessment.

Previous research has concluded that the control and enjoyment one has during social activity could affect the fatigue experienced (Oerlemans & Bakker, 2014; Oerlemans et al., 2014; Ten Brummelhuis & Trougakos, 2014). The significant improvement in social participation restriction (SPARQ) post fitting highlighted a psychosocial benefit of hearing aid fitting, which could in part reflect improved enjoyment or control in social settings. No impact of hearing aid fitting on work or physical activity was evident in the current study.

### Strengths and limitations

The study recruited participants to the intervention group from people who were due to visit an audiology clinic as potential recipients of a hearing aid. As is normal, some of these ended up not being fitted with hearing aids, so they became part of the control group instead. There was a possibility that these participants could respond differently at the second assessment, either with improved scores due to relief (having been told that they did not need hearing aids), or worse scores (having been told they could not be helped at this point). While scores consistent with this behaviour were displayed in some individuals, additional analysis showed that this had no impact on group VFS-AHL scores. Additionally, due to the variable nature of audiology appointments, the time from first assessment to second assessment was longer than three months for some individuals. While this was not a major issue, it could have distorted some analyses.

The difference between groups regarding age range occurred due to the extended period of recruitment and data collection, with some participants finishing as others began. Thus, any dropouts were problematic to match in the opposing group. Finally, most participants had mild to moderate hearing losses, as the study required participants who were only just receiving their first hearing aid. It is possible that hearing aid fitting would result in more dramatic listening-related fatigue reductions for people with severe hearing losses, which this study could not fully investigate. On the other hand, this study represents a realistic sample of people who are likely to receive a first ever hearing aid, and therefore is more clinically relevant than one involving more severely hearing-impaired participants.

The longitudinal design of the study allowed investigation of changes at the individual level across time. By doing so, many of the potential confounding variables in previous research were reduced or eradicated. As the FAS has been used in relevant studies of people with hearing loss before, it was the basis for sample size and power analysis. However, the FAS has, to the authors’ knowledge, never been used in longitudinal assessment before. Despite this, the FAS has been shown to have excellent internal consistency, convergent validity and test-retest reliability for various populations (Hendriks et al., 2018; Horisberger et al., 2019). By using multiple questionnaires to measure fatigue, as well as related variables, the results are more appropriate for understanding the ongoing processes related to the benefit of hearing aid fitting.

## Conclusions

The present study is the first to use longitudinal methodology to measure fatigue before and after first ever hearing aid fitting. It is also the first study to demonstrate a longitudinal increase in social activity level after hearing aid fitting. Hearing aid fitting showed no effect on long-term general fatigue, but there was an improvement from before fitting to six months post fitting for listening-related fatigue. This provides a more detailed result compared to the mixed outcomes from previous research. Like social activity level, social participation restrictions improved post fitting. While the design of this study does not allow an assessment of a direct impact of social activity on fatigue, the increase in social activity and reduction in listening-related fatigue after hearing aid fitting are important findings for clinical practice as more focus should be given to extra-auditory needs and outcomes during auditory healthcare. Future research should assess whether current fatigue scales measure what we want to know in relation to hearing loss and hearing aid fitting. Research should also further investigate the impact of hearing aid use on social activity level, as well as on experiential aspects of social activity such as restriction.

## Supporting information

supplementary digital content 1

supplementary digital content 2

## Data Availability

All data involved in the study is available upon request to the corresponding author.

## Acknowledgments

We would like to thank all participants who took part in the study as well as NHS Greater Glasgow & Clyde Audiology for their help in recruitment. No potential conflict of interest was reported by the authors. This work was supported by the Medical Research Council [grant numbers MR/R502169/1, MR/S003576/1]; and by the Chief Scientist Office of the Scottish Government. The funder had no role in study design.

