## supplementary digital content 1 for "Hearing aids reduce daily-life fatigue and increase social activity: a longitudinal study"

**Self-report outcome measures**

**Fatigue**

**Fatigue Assessment Scale**

The FAS (Michielsen et al., 2003) is a widely used unidimensional general fatigue scale with very good psychometric properties. It has also been used in relevant research involving participants with hearing loss (Alhanbali et al., 2017; Alhanbali et al., 2018). It comprises of 10 questions on a five-point Likert scale. Asking participants how they "usually feel", possible responses are "never", "sometimes", "regularly", "often" and "always". An example question is "I have enough energy for everyday life". The final score range was from 0 to 40, with higher scores indicating greater fatigue.

**Multidimensional Fatigue Symptom Inventory – Short Form**

The MFSI (Stein et al., 2004) has been used primarily in the measurement of cancer-related fatigue, and has been shown to have good psychometric properties for ill and healthy participants (Donovan et al., 2015). It was developed in order to measure separate dimensions of fatigue: general fatigue, physical fatigue, emotional fatigue, mental fatigue and vigour. The questionnaire also provides a total fatigue score by subtracting vigour from the sum of the other scales. Comprising of 30 questions on a five-point Likert scale, participants are asked to rate how true each statement has been of them in the "past seven days". Statements ask about symptoms such as "I feel sluggish". Possible responses are "not at all", "a little", "moderately", "quite a bit" and "extremely". The score range is from -24 to +76, with higher scores indicating greater fatigue.

**Vanderbilt Fatigue Scale – Adult Hearing Loss**

The VFS-AHL (Hornsby et al., 2021) is a valid and reliable tool for measuring listening-related fatigue, encompassing aspects of both transient and long-term fatigue, and comprising of both unidimensional and multidimensional scales. The multidimensional scales assessed in the current study were cognitive fatigue, emotional fatigue and social fatigue. The questionnaire comprises of 40 questions on a five-point Likert scale asking what participants experience in a "typical week". Possible responses are "never/almost never", "rarely", "sometimes", "often" and "always/almost always". An example question is "I become mentally tired when it is hard to listen". The raw scores were transformed using an item response theory scoring algorithm (Hornsby et al., 2021). The resulting final score range for this data was from approximately -2 to 2, with higher scores indicating greater fatigue.

#### Listening effort

##### Listening Effort Assessment Questionnaire (EAS)

The EAS (Alhanbali et al., 2017) measures the amount of effort participants use "listening in everyday life". Responses are given to 6 questions on a visual analogue scale from 0 (No effort) to 10 (lots of effort). The final score range was from 0 to 60, with higher scores indicating greater listening effort.

#### Hearing handicap

##### Hearing Handicap Inventory for the Elderly/Adults (HHIE/A)

The HHIE/A (Ventry & Weinstein, 1982; Newman et al., 1990) consists of two questionnaires aimed at measuring the handicap caused by a hearing loss (HHIE for people aged 65 and over; otherwise HHIA). The questionnaires consist of 25 questions with possible responses of "yes", "sometimes" and "no". Total handicap score was used in this study. The final score range was from 0 to 100, with higher scores indicating greater hearing handicap.

#### Activity

##### Social activity

##### Social Activity Log (SAL)

The SAL (Syrjala et al., 2010) measures the quantity of social activity in the "past week" and "past month". The questionnaire consists of 15 questions asking how many times in the given period the participant undertook different social activities. For most of the questions the possible responses are from '0' to '6 or more'. The final score range was from 0 to 6, with higher scores indicating greater social activity.

##### Social Participation Questionnaire (SPQ)

The SPQ (Densley et al., 2013) is a measure of the quantity of social activity undertaken in the "last twelve months". The questionnaire consists of 22 questions asking how regularly the participant has participated in different social activities. The possible responses on a six point Likert scale are "never", "rarely", "a few times a year", "monthly", "a few times a month" and "once a week or more". In this study only the first 18 questions were used as the final four activities were seldom understood or undertaken by the study sample. The final four questions may be culturally specific. The final score range was from 0 to 90, with higher scores indicating greater social activity.

### Work Activity

#### Craig Handicap Assessment and Reporting Technique (CHART)

The “how do you spend your time?” section of CHART (Whiteneck et al., 1992) was used to measure the work activity of participants. This section of the questionnaire consists of seven questions answered in terms of hours per week spent in different types of paid and unpaid work, with no defined maximum.

### Physical Activity

#### International Physical Activity Questionnaire - Short (IPAQ)

The IPAQ (Craig et al., 2003) was used to measure physical activity in the “last seven days”. The questionnaire gives four scenarios and asks participants to rate on how many days they completed that certain level of exercise (including sitting down) and on average for how long. A total physical activity score is then calculated for the past week with a recommended range of 0 to 19278.

### Social Participation Restrictions

#### Social Participation Restrictions Questionnaire (SPaRQ)

The SPaRQ (Heffernan et al., 2018; Heffernan et al., 2019) consists of two scales assessing ‘social behaviours’ and ‘social perceptions’. The questionnaire is used here as the psychosocial component of social activity. The scales contain nine and ten questions respectively, with possible scores on a visual analogue scale ranging from 0 (completely disagree) to 10 (completely agree). The final score range for each scale was from 0 to 90, and from 0 to 92 respectively, with higher scores indicating greater social participation restriction.

### Auditory Lifestyle

#### Auditory Lifestyle and Demand Questionnaire (ALDQ)

The ALDQ (Gatehouse et al., 1999) consists of twenty-four situations, with participants answering two sub-questions for each. They are asked both “how often” they are in each situation and “how important a factor” the situation is in their everyday life. Possible answers are “very rarely”, “sometimes” and “often”, or “very little”, “some importance” and “very important”. The final score range was from 0 to 100 (frequency x importance), with higher scores indicating a richer auditory environment.

**Need for Cognition**

**Need for Cognition Scale (NFC)**

The NFC (Cacioppo & Petty, 1982) measures the tendency for an individual to engage in and enjoy thinking. The questionnaire consists of 18 statements which prompt the participant to answer how "characteristic" it is of them. Possible answers on a five-point Likert scale are "extremely uncharacteristic", "somewhat uncharacteristic", "uncertain", "somewhat characteristic" and "extremely characteristic". The final score range was from 18 to 90, with higher scores indicating a greater tendency to engage in and enjoy thinking.


Alhanbali, S., Dawes, P., Lloyd, S., & Munro, K. J. (2017). Self-Reported Listening-Related Effort and Fatigue in Hearing-Impaired Adults. *Ear Hear, 38*(1), e39-e48.

Cacioppo, J. T., & Petty, R. E. (1982). The need for cognition. *J Pers Soc Psychol, 42*(1), 116.

Craig, C. L., Marshall, A. L., Sjöström, M., Bauman, A. E., Booth, M. L., Ainsworth, B. E., . . . Sallis, J. F. (2003). International physical activity questionnaire: 12-country reliability and validity. *Medicine & Science in Sports & Exercise, 35*(8), 1381-1395.

Densley, K., Davidson, S., & Gunn, J. M. (2013). Evaluation of the Social Participation Questionnaire in adult patients with depressive symptoms using Rasch analysis. *Quality of Life Research, 22*(8), 1987-1997.

Donovan, K. A., Stein, K. D., Lee, M., Leach, C. R., Ilozumba, O., & Jacobsen, P. B. (2015). Systematic review of the multidimensional fatigue symptom inventory-short form. *Support Care Cancer, 23*(1), 191-212.

Gatehouse, S., Elberling, C., & Naylor, G. (1999). Auditory models and non-linear hearing instruments. *Proc. 18th Danavox Symposium: Auditory models and non-linear hearing instruments*, 221-233.

Heffernan, E., Coulson, N. S., & Ferguson, M. A. (2018). Development of the Social Participation Restrictions Questionnaire (SPaRQ) through consultation with adults with hearing loss, researchers, and clinicians: a content evaluation study. *Int J Audiol, 57*(10), 791-799.

Heffernan, E., Maidment, D. W., Barry, J. G., & Ferguson, M. A. (2019). Refinement and validation of the Social Participation Restrictions Questionnaire: an application of Rasch analysis and traditional psychometric analysis techniques. *Ear Hear, 40*(2), 328-339.

Hornsby, B. W., Camarata, S., Cho, S.-J., Davis, H., McGarrigle, R., & Bess, F. H. (2021). Development and validation of the Vanderbilt Fatigue Scale for Adults (VFS-A). *Psychological Assessment*.

Michielsen, H. J., De Vries, J., & Van Heck, G. L. (2003). Psychometric qualities of a brief self-rated fatigue measure: The Fatigue Assessment Scale. *Journal of Psychosomatic Research, 54*(4), 345-352.

Newman, C. W., Weinstein, B. E., Jacobson, G. P., & Hug, G. A. (1990). The Hearing Handicap Inventory for Adults: psychometric adequacy and audiometric correlates. *Ear Hear, 11*(6), 430-433.

Syrjala, K. L., Stover, A. C., Yi, J. C., Artherholt, S. B., & Abrams, J. R. (2010). Measuring social activities and social function in long-term cancer survivors who received hematopoietic stem cell transplantation. *Psycho-Oncology, 19*(5), 462-471.

Ventry, I. M., & Weinstein, B. E. (1982). The hearing handicap inventory for the elderly: a new tool. *Ear Hear, 3*(3), 128-134.

Whiteneck, G. G., Charlifue, S. W., Gerhart, K. A., Overholser, J. D., & Richardson, G. N. (1992). Quantifying handicap: a new measure of long-term rehabilitation outcomes. *Archives of Physical Medicine and Rehabilitation, 73*(6), 519-526.
