## supplementary digital content 2 for "Hearing aids reduce daily-life fatigue and increase social activity: a longitudinal study"

Figure: Correlation matrix of baseline questionnaires

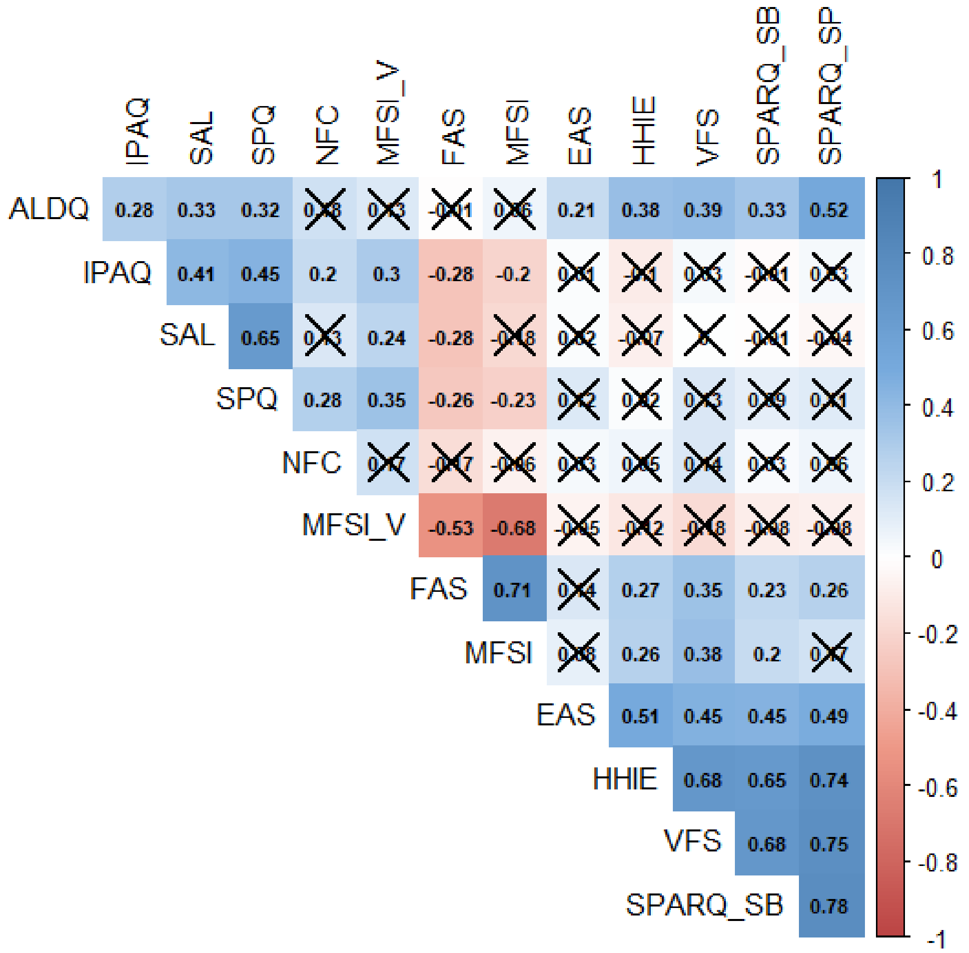


Figure: Correlation matrix of baseline questionnaires. Spearman rank correlation coefficients are colour coded and labelled. Where there was no significant correlation (p > .05), the square is crossed. MFSI_V = MFSI vigour subscale; SPARQ_SB = SPARQ social behaviours subscale; SPARQ_SP = SPARQ social perceptions subscale. Other questionnaire abbreviations can be found in the questionnaires section.

Figure: Individual VFS-AHL score trajectories over time by group

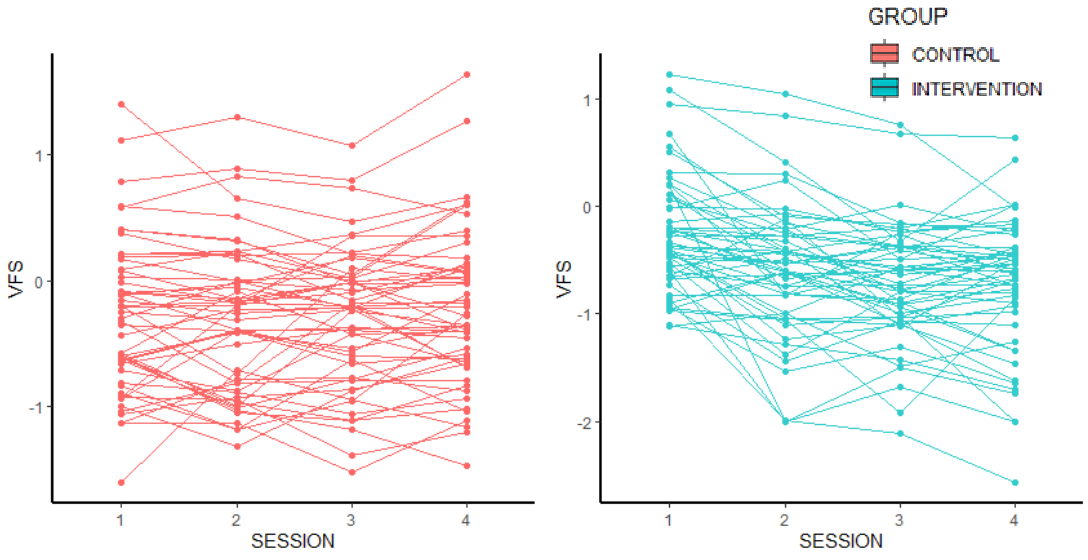

Figure: Individual VFS-AHL score trajectories over time by group. Each line represents one participant's VFS-AHL score from baseline to session four. VFS = Vanderbilt Fatigue Scale – Adult Hearing Loss
